# Comparative adverse effects, perceptions and attitudes related to BNT162b2, mRNA1273, or JNJ-78436735 SARS-CoV-2 vaccines: A population-based longitudinal cohort

**DOI:** 10.1101/2022.09.27.22280403

**Authors:** Oliver Bürzle, Dominik Menges, Julian D. Maier, Daniel Schams, Milo Puhan, Jan Fehr, Tala Ballouz, Anja Frei

## Abstract

**Importance:** Long-term control of SARS-CoV-2 requires effective vaccination strategies. This has been challenged by public mistrust and spread of misinformation regarding vaccine safety. Hence, better understanding and communication on the longer-term and comparative experiences of general population individuals following SARS-CoV-2 vaccination are required.

**Objective:** To evaluate and compare self-reported adverse effects following SARS-CoV-2 vaccination, participants’ perceptions regarding vaccinations and their compliance with recommended public health measures.

**Design, Setting and Participants:** Population-based longitudinal cohort of 575 adults, randomly selected from all individuals presenting to the reference vaccination center of the Canton of Zurich, Switzerland, for receipt of BNT162b2, mRNA1273, or JNJ-78436735.

**Exposures:** BNT162b2, mRNA1273, or JNJ-78436735 vaccines.

**Main Outcomes and Measures:** Primary outcomes included period prevalence, onset, duration, and severity of self-reported adverse effects over 12 weeks following vaccination with a specific focus on the proportion of participants reporting allergic reactions, menstrual irregularities, or cardiac adverse effects, or requiring hospitalization. Secondary outcomes included risk factors associated with reporting adverse effects, perception of vaccine importance, trust in public health authorities and pharmaceutical companies, and compliance with recommended public health measures.

**Results:** 454 (79.0%) participants reported at least one adverse effect during 12 weeks after vaccination. Prevalence was highest among mRNA-1273 recipients (88.7% vs. 77.3% after BNT162b2, 69.1% after JNJ-78436735). Most adverse effects were systemic (72%), occurred within 24 hours (67.9%), and resolved in less than three days (76.3%). 85.2% were reported as mild or moderate. Allergic reactions were reported by 0.4% of participants, hospitalizations by 0.7%, cardiac adverse effects by 1.4%. Menstrual irregularities were reported by 9% of female participants younger than 50 years. Female sex, younger age, higher education, and receipt of mRNA-1273 were associated with reporting adverse effects. Compared to JNJ-78436735 recipients, a higher proportion of mRNA vaccine recipients agreed that vaccination is important (87.5% vs. 28.5%), and trusted public health authorities (80.2% vs. 30.3%) and pharmaceutical companies (71.7% vs. 23.6%).

**Conclusions and Relevance:** Our population-based cohort provided real-world data on self-reported adverse effects following SARS-CoV-2 vaccination and highlights the importance of transparent communication regarding adverse effects and building trust in public health authorities to ensure successful future vaccination campaigns.

**Main Points:** Our representative population-based cohort study demonstrated the safety of three SARS-CoV-2 vaccines and provides real-world estimates on adverse effect incidence.Transparent communication of expected adverse effects to vaccine-seeking individuals is pivotal to build trust in current or future vaccination campaigns.

## Introduction

Beginning with the first vaccinations against SARS-CoV-2 in December 2020, the largest global vaccination campaign in recent history captured the public and professional attention for months. Apart from the obvious focus on efficacy, concerns about vaccine-related adverse effects dominated the professional and public discourse during the campaign. The fast-track authorization of the technologically new mRNA vaccines BNT162b2 (Pfizer-BioNTech) and mRNA1273 (Moderna) in some countries and misinformation contributed to vaccine skepticism and hesitancy.[1-5] This highlights the importance of understanding and accurately communicating information regarding adverse effects, including vaccine safety profiles, to improve vaccine confidence and uptake.

The current body of evidence on adverse effects following SARS-CoV-2 vaccination consists mostly of data reported in randomized clinical trials (RCTs) and reports to government-based surveillance systems, such as the European EudraVigilance, US VAERS (Vaccine Adverse Event Reporting System), or Swiss ElViS (Electronic Vigilance System). Adverse effects reported in RCTs have been primarily mild and self-limited,[6, 7] with systemic reactions (e.g. fatigue, headache, pain) and local injection site reactions (e.g. pain, erythema, swelling) being the most frequent.[8-10] In contrast, severe adverse effects accounted for a significantly higher proportion of reports in governmental surveillance systems.[11-13] This was to be expected as reporting to surveillance systems is subject to several biases including underreporting of mild and common adverse effects and increased reporting of those which are severe or widely reported in the media.[14, 15] Although RCTs provided important evidence on the safety of individual vaccines, they offered little side-by-side comparisons. Furthermore, RCTs yield data collected on selected populations raising the issue of how well this data correlates to “real-world” experiences. The few studies eliciting patient-reported symptoms following different vaccines in real-world settings often had cross-sectional designs and were conducted among very specific groups such as healthcare workers or university students, which may not be representative of the general population.[6, 7] In this population-based study, we aimed to deliver a comprehensive comparative analysis of self-reported adverse effects up to 12 weeks after receipt of three SARS-CoV-2 vaccines approved in Switzerland in 2021. Further objectives were to examine the general perception and attitudes of individuals regarding SARS-CoV-2 vaccination and their compliance with recommended public health measures. Thereby, we aim to improve our understanding of the adverse health effects experienced following vaccination in the general population to provide an evidence base for future vaccination campaigns in view of the likely implementation of additional booster vaccinations and updated SARS-CoV-2 vaccines.

## Methods

### Study design, participants, and recruitment

This study is based on the Zurich SARS-CoV-2 Vaccine Cohort, an ongoing prospective population-based cohort study. We recruited participants between March 10, 2021, and January 27, 2022, at the University of Zurich’s (UZH) vaccination center, the reference center for the Canton of Zurich, Switzerland. All individuals scheduled to receive one of the SARS-CoV-2 vaccines approved in Switzerland in 2021, BNT162b2 (Pfizer-BioNTech), mRNA1273 (Moderna), or JNJ-78436735 (Johnson & Johnson), were screened for eligibility. Eligibility criteria were being 18 years or older, being able to follow study procedures, having sufficient knowledge of the German language and residing in the Canton of Zurich. We excluded individuals who had already received a first dose of a SARS-CoV-2 vaccine. A daily age-stratified (18-64 years, 65 years or older) random sample was selected separately for each approved vaccine from all eligible individuals belonging to the following vaccination groups as defined by the Canton of Zurich [16, 17]: “Over 75 years”, “over 65 years”, “between 50–64 years”, and “between 18–49 years”. We excluded individuals belonging to groups specific for “healthcare workers”, “caretakers of high-risk patients”, “individuals living in communal facilities”, and “individuals with the highest risk diseases” to ensure that that our sample was representative of the general population.[18, 19] Randomly selected individuals were then invited to participate in our study. We obtained written informed consent from all participants. We were unable to reach the desired sample size for JNJ-78436735 recipients 65 years or older, due to limited demand.

The study protocol was prospectively registered on the International Standard Randomized Controlled Trial Number Registry (ISRCTN 15499304) and approved by the ethics committee of the Canton of Zurich (BASEC 2021-00273).

### Data sources and measurements

Upon enrolment, all participants completed a baseline questionnaire including questions on their sociodemographics, medical and smoking history, SARS-CoV-2 related information such as prior infections, perceptions and attitudes regarding vaccination, trust in public health authorities and pharmaceutical companies, and compliance with recommended public health measures (including use of the SwissCovid digital proximity tracing app). Perception, attitude, and compliance questions were collected using a numerical scale (0 “Very low opinion”, to 100 “Very high opinion” for perception), or a 5-item Likert (ranging from “Strongly disagree” to “Strongly agree” for trust-related statements, and from “Never/Impossible” to “Always” for statements on compliance with public health measures). We pooled BNT162b2 and mRNA-1273 recipients into one “mRNA vaccine” group since they expressed similar perceptions towards vaccination and compliance with recommended measures (Supplement 1).

We provided participants with a paper symptom diary and instructed them to record any adverse effect they experienced during 12 weeks after vaccination, as free text, including start and end dates, perceived severity (on a 5-item Likert scale, ranging from “Very mild” to “Very severe”), and consequences of the adverse effects (i.e., self-medication, need for healthcare services, or hospitalizations). Symptom diaries were collected at the 12-week follow-up visit. Participants received additional electronic questionnaires at 4, 6, and 12 weeks, in which they were asked to report any positive SARS-CoV-2 polymerase chain reaction (PCR) or rapid antigen tests. We excluded all reported adverse effects starting within three days before and at any timepoint after positive SARS-CoV-2 tests to ensure that reported symptoms were related to vaccination rather than infection. To determine the proportion of participants with past SARS-CoV-2 infection, we measured participants’ anti-SARS-CoV-2 Spike (S)-IgA and IgG antibodies at baseline using a highly sensitive and specific Luminex technology-based assay.[20]

We collected and managed all study data using the Research Electronic Data Capture (REDCap) system.[21, 22]

### Outcomes

Our primary outcomes included period prevalence, onset, duration, and severity of self-reported adverse effects over 12 weeks following vaccination, with a specific focus on the proportion of participants reporting allergic reactions, menstrual irregularities, or cardiac adverse effects, or requiring hospitalization. Secondary outcomes included risk factors associated with adverse effect reports, general perceptions and attitudes regarding vaccination, trust in public health authorities and pharmaceutical companies, and compliance with recommended public health measures.

### Statistical Analysis

We descriptively analyzed the characteristics and outcomes of interest for the overall cohort and for each of the three vaccine groups. Continuous variables are reported as median with interquartile range (IQR); categorical or ordinal variables as frequencies (N) and percentages (%).We coded adverse effect data reported by participants in the symptom diary according to the Medical Dictionary for Regulatory Activities (MedDRA) hierarchical terminology (Supplement 2).[23] The self-reported adverse effects were translated from German to the closest matching MedDRA “low level term”. All corresponding higher-level terms were included in the database, a highest level of coding was added, labelling each adverse effect either as “local” or “systemic”. We explored associations of several predictor variables on the outcome of reporting one or more adverse effects using a multivariable logistic regression model. Age, sex, vaccine type and education were included a priori variables in the model based on findings from other studies.[6]

Results are presented as odds ratios (OR) with their 95% confidence intervals (CI) and two-sided p-value. All analyses were performed using R (version 4.1.2).

## Results

### Cohort characteristics

Of our 575 participants, 323 participants (56.2%) were female, and the median age was 59 years (IQR 41 to 70) (Table 1). 411 (71.5%) participants received a mRNA-based vaccine (2 doses 3 to 4 eeks apart, 36.2% BNT162b2 and 35.3% mRNA-1273) and 164 (28.5%) received a vector-based vaccine (1 dose, JNJ-78436735). The proportion of participants with a higher level of education (39.8% vs 61.8%) was lower among JNJ-78436735 recipients compared to mRNA vaccine recipients.

**Table 1.** Demographic and clinical characteristics of study population

|  | **BNT162b2**<br>**(Pfizer/BioNTech)** | **mRNA-1273**<br>**(Moderna)** | **JNJ-78436735**<br>**(Johnson & Johnson)** | **Overall** |
| --- | --- | --- | --- | --- |
|  | **(N=207)** | **(N=203)** | **(N=165)** | **(N=575)** |
| **Age, median (IQR) -in years** | 59 (36 to 75) | 65 (42.5 to 69) | 58 (45 to 70) | 59 (41 to 70) |
| **Age distribution - no. of participants (%)** |  |  |  |  |
| <65 years | 106 (51.2%) | 99 (48.8%) | 103 (62.4%) | 308 (53.6%) |
| ≥65 years | 101 (48.8%) | 104 (51.2%) | 62 (37.6%) | 267 (46.4%) |
| **Female sex - no. of participants (%)** | 115 (55.6%) | 120 (59.1%) | 88 (53.3%) | 323 (56.2%) |
| **Presence of at least one preexisting medical condition (%)** | 80 (40.8%) | 78 (42.2%) | 64 (43.2%) | 222 (42.0%) |
| **Highest educational level - no. of participants (%)** |  |  |  |  |
| None or mandatory school | 9 (4.4%) | 4 (2.0%) | 7 (4.3%) | 20 (3.5%) |
| Vocational training or specialized<br>baccalaureate | 71 (34.5%) | 72 (35.6%) | 91 (55.8%) | 234 (41.0%) |
| Higher technical school or college | 46 (22.3%) | 44 (21.8%) | 40 (24.5%) | 130 (22.8%) |
| University | 80 (38.8%) | 82 (40.6%) | 25 (15.3%) | 187 (32.7%) |
| Missing | 1 (0.5%) | 1 (0.5%) | 2 (1.2%) | 4 (0.7%) |
| **Reported adverse effects - no. of participants (%)** | 160 (77.3%) | 180 (88.7%) | 114 (69.1%) | 456 (79.3%) |
| **Tested seropositive for anti-SARS-CoV-2 S-IgA at baseline - no. of participants (%)** | 12 (5.8%) | 14 (6.9%) | 20 (12.1%) | 46 (8.0%) |
| **Tested seropositive for anti-SARS-CoV-2 S-IgG at baseline - no. of participants (%)** | 15 (7.2%) | 19 (9.4%) | 24 (14.5%) | 58 (10.1%) |
| **Reported positive SARS-CoV-2 test prior to baseline - no. of participants (%)** | 8 (3.9%) | 13 (6.4%) | 16 (9.7%) | 37 (6.4%) |
| **SARS-CoV-2 Infection prior to baseline (self-reported infection or tested seropositive)- no. of participants (%)** | 21 (10.1%) | 28 (13.8%) | 31 (18.8%) | 80 (13.9%) |
| **Tested positive for anti-SARS-CoV-2 S-IgA or IgG at baseline with no report of prior SARS-CoV-2 Infection- no. of participants (%)** | 13 (6.3%) | 15 (7.4%) | 15 (9.1%) | 43 (7.5%) |

37 participants (6.4%) reported ever having a positive SARS-CoV-2 test prior to vaccination, with a higher proportion among JNJ-78436735 recipients (9.7% vs 6.4% of mRNA-1273 and 3.7% of BNT162b2 recipients). 19 (9.2%) BNT162b2 recipients, 24 (11.8%) mRNA-1273 recipients and 29 (17.6%) JNJ-78436735 recipients tested seropositive for anti-SARS-CoV-2 S-IgA or -IgG before receiving the first vaccination dose.

### Frequency and characteristics of adverse effects

Overall, 79.0% (N= 454) of all participants reported at least one adverse effect up to three months following vaccination, with a total of 2397 reported adverse effects. The highest proportion of participants with adverse effects after vaccination was among mRNA-1273 recipients (88.7%, N=180) compared to BNT162b2 (77.3%, N=160) and JNJ-78436735 (69.1%, N=114) recipients. Based on a multivariable logistic regression model, we found strong to very strong evidence that female sex (OR=4.05 (95% CI: 2.33 to 7.3), p<0.001), higher education levels (vs. none or mandatory school, OR=6.26 (1.86 to 21.2), p=0.003) and receiving mRNA-1273 (vs. BNT162b2, OR=2.38 (1.22 to 4.8), p=0.013) were associated with adverse effect reports. There was weak evidence that younger age (<65 vs. ≥65 years, OR=1.65 (0.96 to 2.9), p=0.072) was associated with adverse effects. We found no evidence that JNJ-78436735 (vs. BNT162b2), preexisting conditions, low opinion (opinion value <50) about vaccination and SARS-CoV-2 infections prior to vaccination were associated with adverse effects (Supplement 3).

More participants reported systemic (71.7%, N=412) than local adverse effects (54.8%, N=315). Among mRNA vaccine recipients, the proportion of systemic among all adverse effects increased after the 2^nd^ dose (63.9% to 77.2% in BNT162b2 and 59.1% to 78.0% in mRNA-1273 recipients, Supplement 4). The most common adverse effect mentioned by mRNA vaccine recipients was local pain (54.1% of BNT162b2 and 69.5% of mRNA-1273 recipients), followed by asthenia (fatigue; 38.7% of BNT162b2 and 44.8% of mRNA-1273 recipients). JNJ-78436735 recipients most frequently reported headache (36.4%), followed by local pain (30.9%) and asthenia (30.9%). Other commonly reported adverse effects included nausea, vertigo, and sore throat (all >5%). Of our participants, 0.4% (n=2) (one BNT162b2 and one mRNA-1273 recipient) reported allergic reactions. Adverse effects affecting menstruation were reported by 5 out of 47 (10.6%) female participants younger than 50 among BNT162b2 recipients, 4 out of 42 (9.5%) among mRNA-1273 recipients and 2 out of 31 (6.5%) among JNJ-78436735 recipients (six participants reported cycle irregularities, three heavy menstrual bleeding, three intermenstrual bleeding). Tachycardia or palpitations were reported by seven (1.2%) participants, four mRNA-1273, two JNJ-78436735 and one BNT162b2 recipient. One BNT162b2 recipient reported pericardial effusion and atrial fibrillation after the second dose.

Most adverse effects (83.9%) occurred in the first week following vaccination, 67.9% within 24 hours. Participants reported that adverse effects lasted for 3.9 days on average, and most resolved within 3 (76.3%) days. Asthenia, extremity pain, and cough were most frequently reported to last longer than a week. Adverse effect onset and duration were similar across the three vaccines (Figure 2A, Supplement 5).

**Figure 1.**
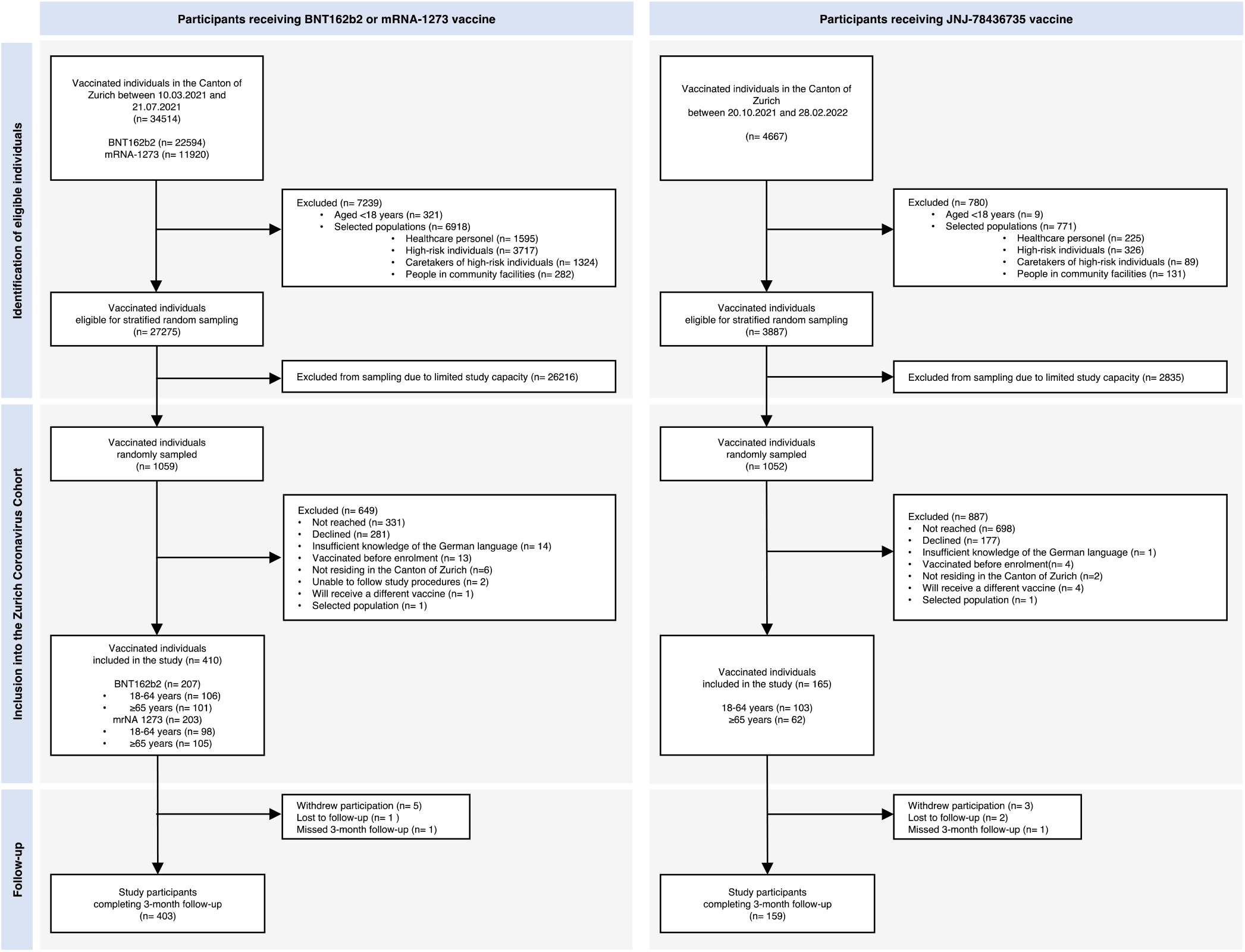
Recruitment of study cohort

**Figure 2.**
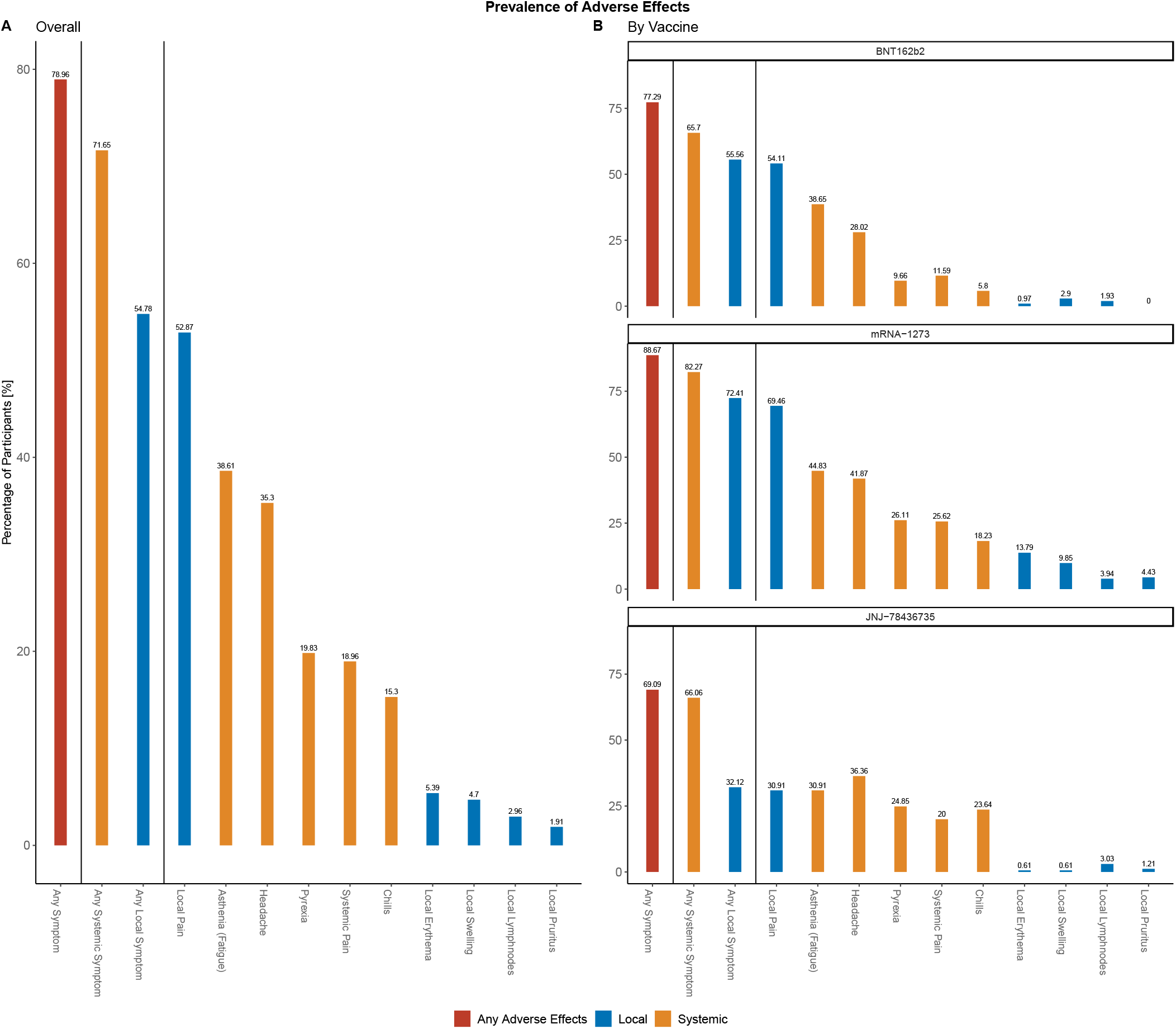
Frequency of any, local and systemic adverse effects. Panel A shows five most common systemic and local adverse effects in overall sample (N=575). Panel B shows adverse effects by vaccine type (BNT162b2 N=208, mRNA-1273 N=203, JNJ-78436735 N=164).

The perceived severity of most adverse effects was mild (49.0%) or moderate (36.2%). Meanwhile, 14.7% were described as severe (13.4%) or very severe (1.3%), with the highest proportion of severe to very severe adverse effects reported after JNJ-78436735 (18.3% vs 16.1% after mRNA-1273, 9.4% after BNT162b2). Asthenia (13.1%), headache (12.8%) and pain (9.4%) were mostly reported as severe or very severe adverse effects. Hospitalization due to reported adverse effects was reported by 0.7% (n=4) of participants (two BNT162b2 recipients with loss of consciousness and bullous pemphigoid, one mRNA-1273 recipient with retinal detachment and one JNJ-78436735 recipient with meningitis).

Most reported adverse effects resolved spontaneously (Figure 2C). However, participants reported using self-prescribed medications (e.g., Paracetamol or Ibuprofen) or seeking consultation with a healthcare provider for 448 (18.7%) and 84 (3.5%) of the adverse effects, respectively.

### Perceptions of vaccination and compliance with recommended public health measures

More mRNA vaccine recipients (87.5%) agreed completely or in part with the statement that it was important to be vaccinated compared to 28.5% of JNJ-78436735 recipients. Similarly, more mRNA vaccine recipients felt that vaccines were a part of a healthy lifestyle (63.6% vs. 28.9% of JNJ-78436735 recipients). Trust in public health authorities (80.2% vs. 30.3%) and pharmaceutical companies (71.7% vs. 23.6%) was higher among mRNA vaccine recipients compared to JNJ-78436735 recipients. Both groups felt they had sufficient understanding of how the vaccine helped the body fend off infectious diseases (89.3% mRNA vaccine recipients vs. 62.4% JNJ-78436735 recipients) and reported similar compliance with recommended public health measures (Figure 3B). Use of the SwissCovid digital proximity tracing app was higher among mRNA vaccine recipients compared to JNJ-78436735 recipients (53.2% vs 27.4%).

**Figure 3.**
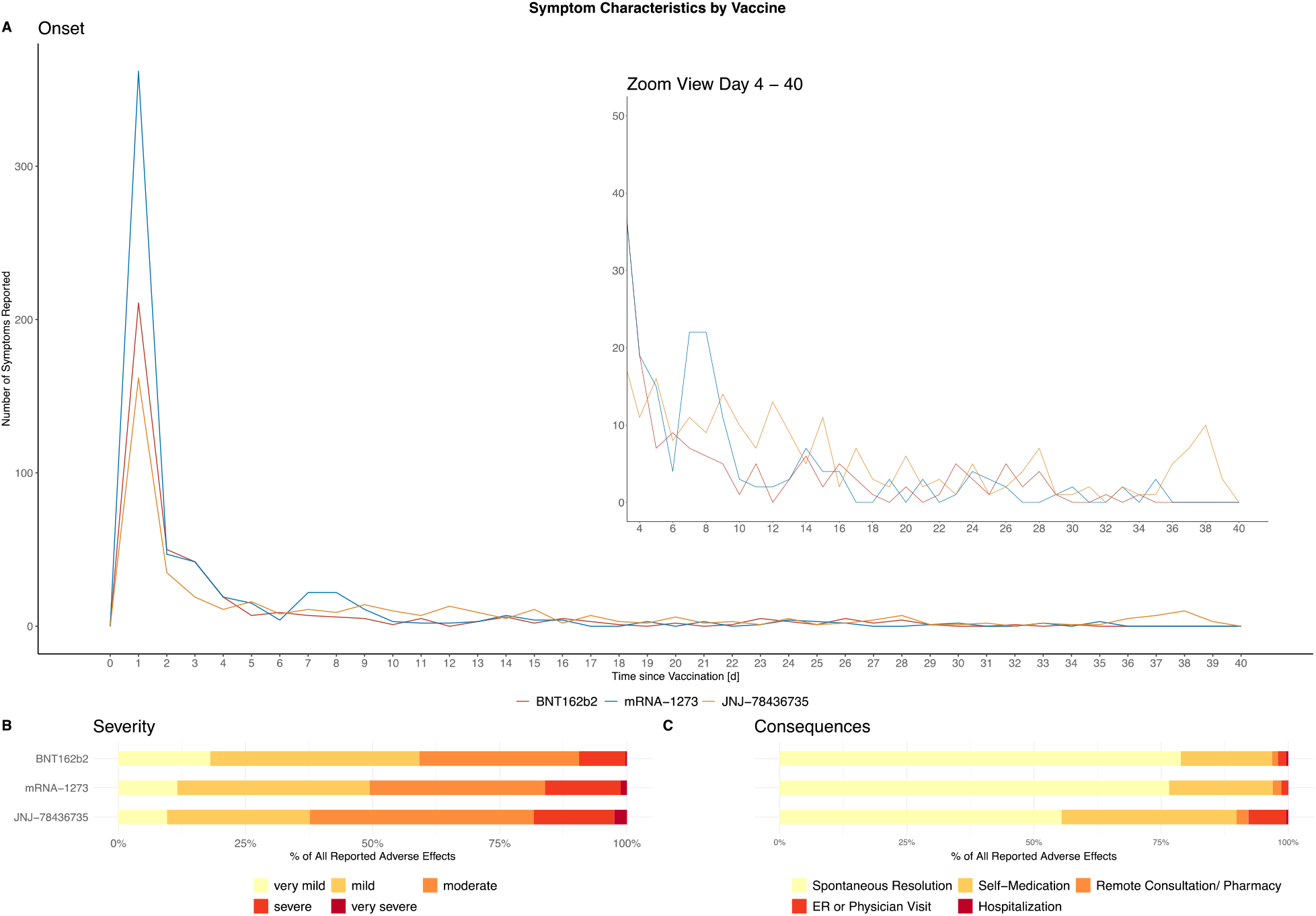
Characteristics of self-reported adverse effects. Panel A shows the time of adverse effect onset by vaccine, panel B perceived adverse effect severity by vaccine and panel C the consequences of adverse effects by vaccine. (ER: Emergency Room)

**Figure 4.**
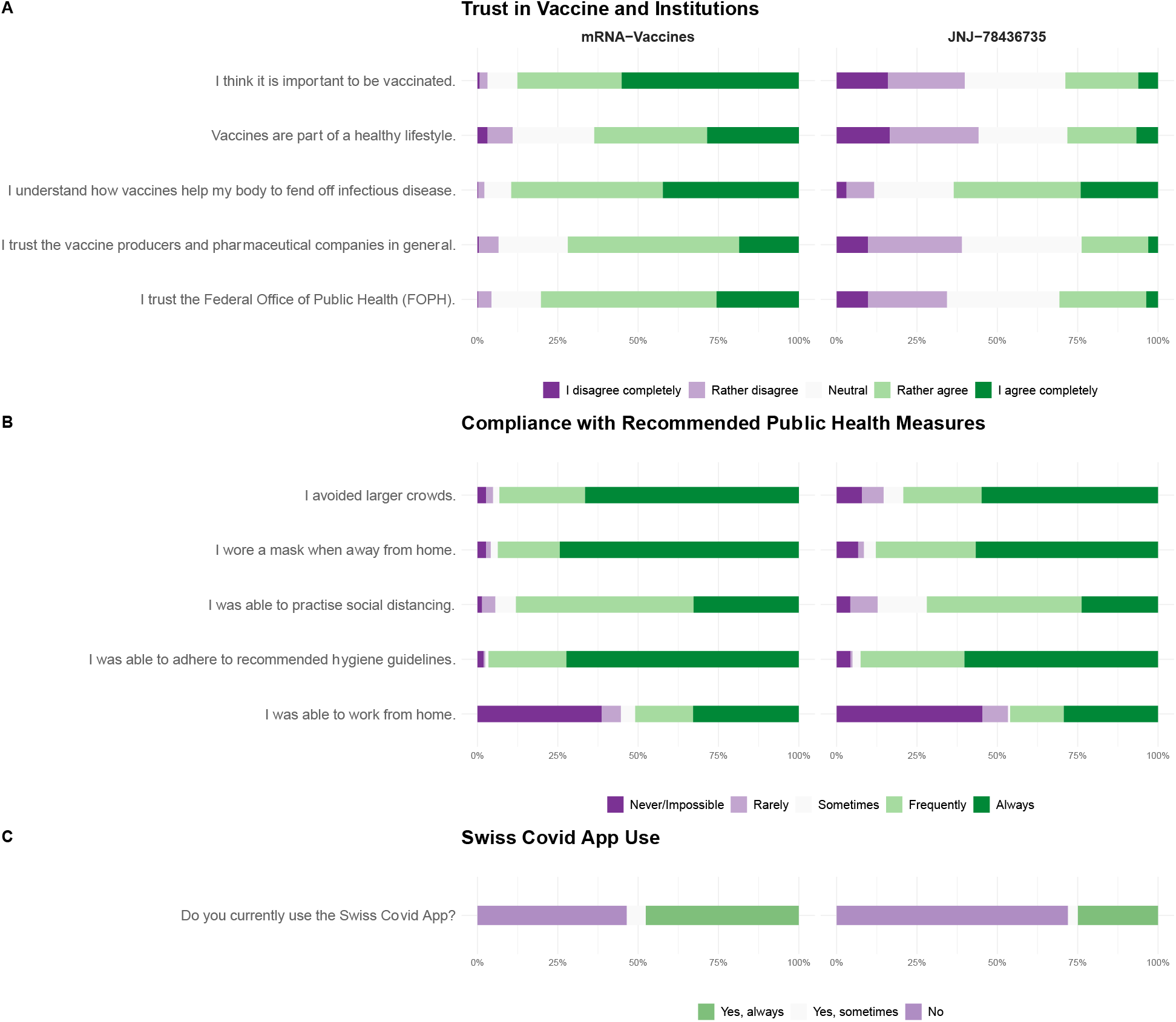
Perception of vaccination and compliance with recommended public health measures.

## Discussion

In this population-based cohort of 575 individuals who received a SARS-CoV-2 vaccine and were followed-up over 12 weeks, participants commonly reported adverse effects, namely local pain, fatigue, headache, and fever. Most adverse effects were mild to moderate and resolved within three days. Allergic reactions (0.4%) and adverse effects requiring hospitalization (0.7%) were rare. Around 9% of female participants younger than 50 reported menstrual cycle changes, more frequently among mRNA vaccine recipients. Female sex, receiving mRNA-1273, higher education and younger age were associated with experiencing adverse effects. JNJ-78436735 recipients less frequently perceived vaccination to be important and had lower trust in public health authorities and pharmaceutical companies compared to mRNA vaccine recipients. There were no differences between vaccine groups in compliance with preventive public health measures.

Our results on the prevalence and severity of adverse effects are in line with previously reported data from RCTs and other observational studies.[6, 9, 10, 24-26] In an online survey among individuals vaccinated with either BNT162b2, mRNA-1273, or JNJ-78436735, Beatty et al. reported that 80.3% of participants experienced adverse effects, with comparable estimates for each vaccine type.[6] Our data also matches the prevalence published in the RCTs for each vaccine individually.[8-10]. The proportion of adverse effects that were self-assessed to be severe or required hospitalization in our study (14.8%) was well below that of Swiss and European governmental surveillance systems (37.9% in Swiss ElViS).[11, 12] US surveillance reports also stated higher estimates of serious adverse events based on hospitalization rates, serious illness and deaths (9.2% vs. our 0.7%).[13] These higher estimates from governmental reporting systems are likely related to the underreporting of mild symptoms and underscore the importance of “real-world” data.[14, 15] There is wide variation in reports on prevalence of anaphylaxis and severe allergic reactions ranging from 0.03% to 3%, due to differing definitions.[6, 27, 28] In our study, two (0.4%) participants reported allergic reactions, without requiring medical attention.

Our follow-up over 12 weeks allowed us to assess adverse effects that occur with some delay, such as menstrual changes reported in 9% of female participants younger than 50 years. Few studies have described menstrual irregularities following SARS-CoV-2 vaccination with prevalence ranging between 0.3% and 46%.[29, 30] This large variability and the high prevalence (37.8%) of menstrual irregularities in the general population regardless of vaccination underscore the challenge of attributing changes in the menstrual cycle to vaccination.[31] Further research is needed on the influence of SARS-CoV-2 vaccination on menstruation and the general impact of vaccination on female recipients, as we and others observed that female recipients were generally more likely to experience adverse effects.[32-35]

We also found that participants who were younger than 65 years and received mRNA-1273 were more likely to report adverse effects possibly due to stronger immune responses among these groups.[6, 13, 26] Similarly, other studies have found evidence that SARS-CoV-2 infections prior to vaccination may be associated with adverse effects reports due to increased immunogenicity.[6, 7] While we found that individuals with prior SARS-CoV-2 infections were 1.8 times more likely to report adverse effects compared to those without, the findings were not statistically significant, potentially due to insufficient power in our study.

mRNA vaccine recipients trusted vaccines in general and thus were mostly motivated to be vaccinated as soon as SARS-CoV-2 vaccines became available. JNJ-78436735 recipients were more hesitant and waited for JNJ-78436735’s introduction to Switzerland, resulting in a higher proportion of individuals with infections prior to vaccination compared to mRNA vaccine recipients. Other studies describe these concerns about the rapid development and fears of adverse effects to be among the main reasons to wait for non-mRNA-based vaccines or other alternatives.[36-38] General skepticism and the presence of nocebo effects, as demonstrated by a Amanzio et al, may have translated into a higher proportion of JNJ-78436735 recipients perceiving adverse effects as severe. [38-40] Increasing awareness of these nocebo responses and using positive framing around the low risk of severe adverse effects may contribute to improving vaccine acceptance.

This study provided evidence from a representative cohort recruited from the general population and followed-up over a 12-week period. Data collected through symptom diaries and adverse effect coding according to MedDRA terms generated a comprehensive dataset allowing a comparative analysis of three common SARS-CoV-2 vaccines.

However, there are several limitations to our study. First, self-selection bias may have occurred if individuals who are more health literate or less hesitant participated in our study, leading to overestimations of trust in public health authorities and more positive perceptions of vaccines. However, we consider the data on the prevalence of adverse effects as broadly representative. Second, our data is self-reported. While this allows for an accurate description of vaccine recipients’ experiences, it is subjective and no verification of the relation of adverse effects and vaccination by a healthcare provider was possible. Third, the absolute numbers of reports for some adverse effects when analyzed individually are relatively small (e.g., menstrual changes). Finally, our analysis was restricted to basic immunization of the three SARS-CoV-2 vaccines approved in Switzerland at the time of study conduct. Further research on adverse effects occurring after booster vaccinations, other types of vaccines and combinations of different vaccines is needed.

## Conclusion

This study demonstrates the safety of three SARS-CoV-2 vaccines in a representative population-based cohort and provides real-world estimates on adverse effect prevalence after vaccination. Thereby, we importantly extend the evidence base for health-care providers to answer many of the questions of individuals seeking vaccination. While further evidence on adverse effects after booster vaccination and other vaccine types is required, our study suggests that transparent communication regarding adverse effects and building trust in public health authorities are pivotal future vaccination campaigns’ success.

## Data Availability

All data produced in the present study are available upon reasonable request to the authors.

## Acknowledgements

The authors thank the study administration team and the team from the University of Zurich’s (UZH) vaccination center, the reference center for the Canton of Zurich, Switzerland, for their dedicated support of the study. They also thank the study participants for their valuable contribution to this project.

