## Supplemental Material for "Comparative adverse effects, perceptions and attitudes related to BNT162b2, mRNA1273, or JNJ-78436735 SARS-CoV-2 vaccines: A population-based longitudinal cohort"

### Supplementary Material

**Supplement 1.** Answers to trust related questions among mRNA vaccine recipients

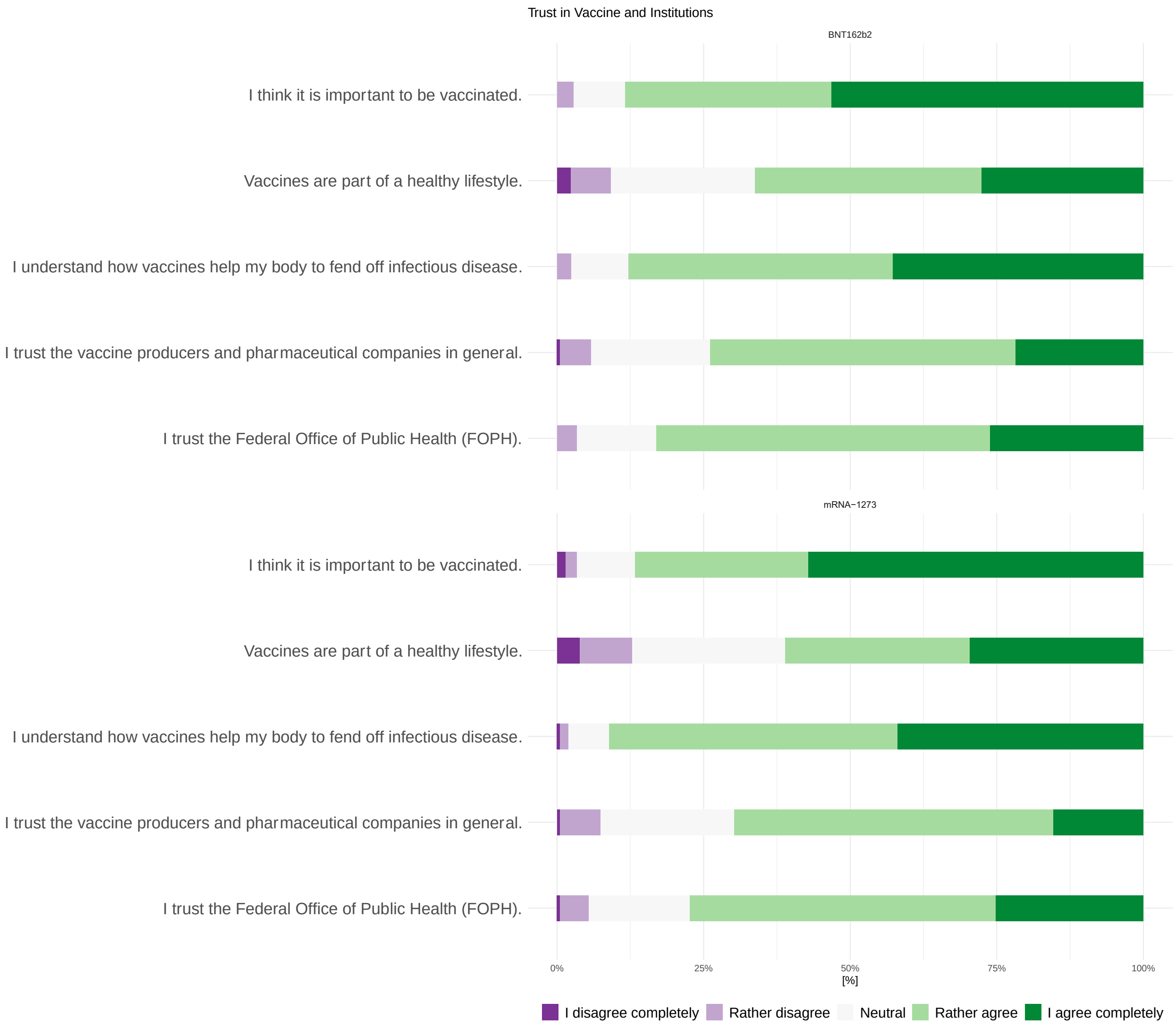

**Supplement 2**. MedDRA coding hierarchy (NEC= not elsewhere classified)

| System Organ Class | High Level Group Term | High Level Term | Preferred Term | Low Level Term |
|---|---|---|---|---|
| blood and lymphatic system disorders | spleen, lymphatic and reticuloendothelial system disorders | lymphatic system disorders NEC | lymphadenopathy | axillary lymph nodes enlarged |
| | | | | lymph nodes cervical swollen |
| | | | | lymph nodes enlarged |
| cardiac disorders | cardiac arrhythmias | supraventricular arrhythmias | atrial fibrillation | atrial fibrillation |
| | | rate and rhythm disorders NEC | tachycardia | tachycardia |
| | cardiac disorders, signs, and symptoms NEC | cardiac signs and symptoms NEC | palpitations | palpitations |
| | pericardial disorders | pericardial disorders NEC | pericardial effusion | pericardial effusion |
| ear and labyrinth disorders | aural disorders NEC | ear disorders NEC | ear inflammation | ear inflammation |
| | | | ear pain | ear pain |
| eye disorders | eye disorders NEC | lacrimation disorders | dry eye | dry eyes |
| | | | lacrimation increased | watering eyes |
| | | ocular disorders NEC | ocular discomfort | sensation of pressure in eye |

| System Organ Class | High Level Group Term | High Level Term | Preferred Term | Low Level Term |
|---|---|---|---|---|
| | ocular infections, irritations and inflammations | conjunctival infections, irritations and inflammations | conjunctivitis | conjunctivitis |
| | | lid, lash and lacrimal infections, irritations and inflammations | blepharitis | blepharitis |
| | | ocular infections, inflammations and associated manifestations | eye irritations | burning eyes |
| | | | ocular hyperemia | eye red |
| | ocular structural change, deposit and degeneration NEC | retinal structural change, deposit and degeneration | retinal detachment | retinal detachment |
| | vision disorders | visual disorders NEC | vision blurred | blurred vision |
| | | visual impairment and blindness (excluding color blindness) | visual impairment | visual impairment |
| gastrointestinal disorders | dental and gingival conditions | gingival Infections | gingivitis | gingivitis |
| | | gingival disorders, signs and symptoms NEC | Non-infective gingivitis | sores gum |
| | | dental pain and sensation disorders | toothache | tooth pain |

| System Organ Class | High Level Group Term | High Level Term | Preferred Term | Low Level Term |
|---|---|---|---|---|
| | gastrointestinal infections | gastric and gastrointestinal infections | gastroenteritis viral | stomach flu |
| | gastrointestinal motility and defecation conditions | diarrhea (excluding infective) | diarrhea | diarrhea |
| | gastrointestinal signs and symptoms | dyspeptic signs and symptoms | dyspepsia | digestion impaired |
| | | | | stomach burning sensation |
| | | flatulence, bloating and distension | abdominal distension | abdominal bloating |
| | | | flatulence | flatulence |
| | | gastrointestinal and abdominal pains (excluding oral and throat) | abdominal pain | abdominal cramps |
| | | | | abdominal pain |
| | | | abdominal pain upper | stomachache |
| | | | | stomach cramps |
| | | | | stomach pain |
| | | gastrointestinal signs and symptoms NEC | abdominal discomfort | abdominal discomfort |
| | | | | stomach discomfort |
| | | | breath odor | bad breath |

| System Organ Class | High Level Group Term | High Level Term | Preferred Term | Low Level Term |
| --- | --- | --- | --- | --- |
| | | | odynophagia | swallowing painful |
| | | nausea and vomiting symptoms | nausea | nausea |
| | | | vomiting | vomiting |
| | gastrointestinal vascular conditions | hemorrhoids and gastrointestinal varices (excluding oesophageal) | haemorrhoidal haemorrhage | haemorrhoidal bleeding |
| | oral soft tissue conditions | oral soft tissue signs and symptoms | hypoaesthesia oral | numbness of tongue |
| | | | lip discoloration | lip discolouration |
| | | | oral pain | sensitive mouth |
| | saliva gland conditions | oral dryness and saliva altered | dry mouth | dry mouth |
| | tongue conditions | tongue signs and symptoms | tongue discolouration | tongue white |
| general disorders and administration site conditions | administration site reactions | injection site reactions | injection site erythema | injection site redness |
| | | | injection site pruritus | injection site itching |
| | | vaccination site reactions | vaccination site discolouration | vaccination site discolouration |
| | | | vaccination site haematoma | vaccination site hematoma |

| System Organ Class | High Level Group Term | High Level Term | Preferred Term | Low Level Term |
|---|---|---|---|---|
| | | | vaccination site inflammation | vaccination site inflammation |
| | | | vaccination site irritation | vaccination site irritation |
| | | | vaccination site movement impairment | vaccination site movement impairment |
| | | | vaccination site pain | vaccination site pain |
| | | | vaccination site rash | vaccination site rash |
| | | | vaccination site swelling | vaccination site swelling |
| | | | vaccination site tenderness | vaccination site tenderness |
| | | | vaccination site warmth | vaccination site warmth |
| | body temperature conditions | febrile disorders | pyrexia | fever |
| | | | | feverish |
| | general system disorders NEC | asthenic conditions | asthenia | energy decreased |
| | | | | fatigue |
| | | | | feeling of weakness |

| System Organ Class | High Level Group Term | High Level Term | Preferred Term | Low Level Term |
|---|---|---|---|---|
| | | | | feelings of weakness |
| | | | | weakness |
| | | | listlessness | listlessness |
| | | | malaise | feeling unwell |
| | | | | unwell |
| | | feelings and sensations NEC | chills | chills |
| | | | decreased appetite | appetite absent |
| | | | feeling abnormal | feeling dazed |
| | | | feeling cold | feeling cold |
| | | | feeling hot | feeling hot |
| | | | | feeling of warmth |
| | | | | sensation of heat |
| | | | hot flush | feeling of hot flushes |
| | | | hunger | feeling hungry |
| | | | irritability | feeling irritated |
| | | | peripheral coldness | cold extremities |
| | | | | cold feet |
| | | | temperature intolerance | heat sensitivity |
| | | | thirst | thirst |
| | | | balance disorder | unsteadiness |

| System Organ Class | High Level Group Term | High Level Term | Preferred Term | Low Level Term |
|---|---|---|---|---|
| | | general signs and symptoms NEC | hot flushes | hot flashes |
| | | | | hot flushes |
| | | | hyperhidrosis | excess sweating |
| | | | | heavy sweating |
| | | | | sweating |
| | | | | sweating attack |
| | | | influenza like illness | flu-like symptoms |
| | | | night sweats | night sweats |
| | | | peripheral swelling | swelling arm |
| | | | | swelling of legs |
| | | | swelling | swelling |
| | | oedema NEC | oedema peripheral | leg edema |
| | | pain and discomfort NEC | axillary pain | armpit pain |
| | | | chest discomfort | chest pressure |
| | | | chest pain | chest burning |
| | | | | chest pain |
| | | | | thorax pain |
| | | | pain | general body pain |
| | | | | pain |
| immune system disorders | allergic conditions | allergic conditions NEC | hypersensitivity | allergic reaction |
| | | angioedemas | swollen tongue | swollen tongue |

| System Organ Class | High Level Group Term | High Level Term | Preferred Term | Low Level Term |
| --- | --- | --- | --- | --- |
| | | urticarias | urticaria | urticaria |
| infections and infestations | viral infectious disorders | herpes viral infections | oral herpes | cold sores |
| | | | | cold sores lip |
| | | | | herpes labialis |
| investigations | cardiac and vascular investigations (excl enzyme tests) | heart rate and pulse investigations | heart rate increased | pulse rate increased |
| | physical examination and organ system status topics | physical examination procedures and organ system status | body temperature increased | temperature elevation |
| metabolism and nutrition disorders | electrolyte and fluid balance conditions | total fluid volume increase | oedema | oedematous weight gain |
| musculoskeletal and connective tissue disorders | bone disorders (excl congenital and fractures) | bone related signs and symptoms | bone pain | bone pain |
| | | bone disorders NEC | exostosis | bone spur |
| | joint disorders | joint related signs and symptoms | arthralgia | joint pain |
| | | | | knee pain |
| | | | | pain in joint |
| | | | | pain in joint involving hand |
| | | | | painful joints |
| | | | | shoulder pain |

| System Organ Class | High Level Group Term | High Level Term | Preferred Term | Low Level Term |
|---|---|---|---|---|
| | | osteoarthropathies | osteoarthritis | arthrosis |
| | | | | gonarthrosis |
| | muscle disorders | muscle pains | myalgia | muscle pain |
| | | | | muscle soreness |
| | | | | tenderness muscle |
| | | muscle related signs and symptoms NEC | muscle spasms | leg cramps |
| | | | | muscle cramps |
| | | | muscle swelling | muscle swelling |
| | | | muscle twitching | muscle twitching |
| | | muscle weakness conditions | muscular weakness | muscle weakness lower limb |
| | | myopathies | myopathy | myopathy |
| | musculoskeletal and connective tissue disorders NEC | musculoskeletal and connective tissue conditions NEC | musculoskeletal stiffness | neck stiffness |
| | | | | stiffness shoulder |
| | | musculoskeletal and connective tissue pain and discomfort | back pain | back pain |
| | | | | lumbago |
| | | | | muscular back pain |
| | | | flank pain | flank pain |
| | | | limb discomfort | feeling heavy in arms and legs |

| System Organ Class | High Level Group Term | High Level Term | Preferred Term | Low Level Term |
|---|---|---|---|---|
| | | | | heavy feeling in arms and legs |
| | | | musculoskeletal pain | pain neck/ shoulder |
| | | | neck pain | neck pain |
| | | | pain in extremity | leg pain |
| | | | | pain foot |
| | | | | pain in fingers |
| | | | | pain in thumb |
| | | | | pain in toe |
| | | | | painful arm |
| | | | | painful feet |
| | | | | painful hand |
| | tendon, ligament and cartilage disorders | tendon disorders | tenosynovitis | tendovaginitis |
| nervous system disorders | central nervous system infections and inflammations | meningitis NEC | meningitis | meningitis |
| | cranial nerve disorders (excl neoplasms) | olfactory nerve disorders | anosmia | smell loss |
| | | auditory nerve disorders | tinnitus | subjective tinnitus |
| | headaches | headaches NEC | headache | headache |

| System Organ Class | High Level Group Term | High Level Term | Preferred Term | Low Level Term |
|---|---|---|---|---|
| | | | | throbbing headache |
| | | migraine headaches | migraine | migraine |
| | | | | migraine with aura |
| | movement disorders (incl parkinsonism) | tremor (excl congenital) | tremor | shaking of hands |
| | neurological disorders NEC | disturbances in consciousness NEC | loss of consciousness | Consciousness loss of |
| | | neurological signs and symptoms NEC | head discomfort | head pressure |
| | | paraesthesia and dysaesthesia | paraesthesia | tingling sensation |
| | | sensory abnormalities NEC | sensory loss | loss of sensation |
| | | vertigos NEC | vertigo | vertigo |
| psychiatric disorders | cognitive and attention disorders and disturbances | cognitive and attention disorders and disturbances NEC | disturbance in attention | poor concentration |
| | deliria (incl confusion) | confusion and disorientation | confusional state | confusion |
| | dementia and amnestic conditions | amnestic symptoms | memory impairment | forgetfulness |

| System Organ Class | High Level Group Term | High Level Term | Preferred Term | Low Level Term |
|---|---|---|---|---|
| | depressed mood disorders and disturbances | depressive disorders | depression | depression |
| | | mood alterations with depressive symptoms | depressed mood | depressed mood |
| | mood disorders and disturbances NEC | emotional and mood disturbances NEC | euphoric mood | euphoria |
| | sleep disorders and disturbances | disturbances in initiating and maintaining sleep | insomnia | sleeplessness |
| | | dyssomnias | poor quality sleep | poor sleep |
| | | | | sleep restless |
| | | sleep disorder NEC | sleep disorder | disorder sleep |
| | | | | sleep problem |
| | somatic symptom and related disorders | somatic symptom disorders | conversion disorder | Functional neurological symptom disorder |
| renal and urinary disorders | bladder and bladder neck disorders (excl calculi) | bladder disorders NEC | bladder disorder | bladder disorder |
| | | bladder infections and inflammations | cystitis haemorrhagic | cystitis hemorrhagic |
| | urinary tract signs and symptoms | bladder and urethral symptoms | dysuria | painful urination |
| | | | incontinence | incontinence |
| | | | micturition urgency | urgency urination |

| System Organ Class | High Level Group Term | High Level Term | Preferred Term | Low Level Term |
|---|---|---|---|---|
| reproductive system and breast disorders | breast disorders | breast signs and symptoms | breast pain | mastodynia |
| | menstrual cycle and uterine bleeding disorders | menstrual and uterine bleeding NEC | intermenstrual bleeding | spotting between menses |
| | | menstruation with increased bleeding | heavy menstrual bleeding | heavy menstrual bleeding |
| | | | menstruation irregular | menstrual irregularity |
| | | | | menstruation irregular |
| | vulvovaginal disorders (excl infections and inflammations) | vulvovaginal disorders NEC | vaginal haemorrhage | spotting vaginal |
| respiratory, thoracic and mediastinal disorders | pleural disorders | pneumothorax and pleural effusions NEC | pleural effusion | pleural effusion |
| | respiratory disorders NEC | breathing abnormalities | dyspnoea | difficulty breathing |
| | | coughing and associated symptoms | cough | cough |
| | respiratory tract infections | upper respiratory tract infections NEC | laryngitis | laryngitis |
| | | | nasopharyngitis | cold symptoms |
| | | | sinusitis | sinusitis |

| System Organ Class | High Level Group Term | High Level Term | Preferred Term | Low Level Term |
|---|---|---|---|---|
| | | viral upper respiratory tract infections | influenza | flu symptoms |
| | respiratory tract signs and symptoms | lower respiratory tract signs and symptoms | hiccups | hiccups |
| | | upper respiratory signs and symptoms | aphonia | loss of voice |
| | | | dry throat | dry throat |
| | | | speech disorder | disorder speech |
| | | upper respiratory tract signs and symptoms | oropharyngeal pain | sore throat |
| | | | rhinorrhoea | rhinorrhea |
| | | | sneezing | sneezing |
| | upper respiratory tract disorders (excl inflammation) | nasal congestions and inflammation | nasal congestion | nose congestion |
| | | | rhinitis allergic | allergic rhinitis |
| | | nasal disorders NEC | epistaxis | nosebleed |
| skin and subcutaneous tissue disorders | epidermal and dermal conditions | bullous conditions | pemphigoid | bullous pemphigoid |
| | | dermal and epidermal conditions NEC | dry skin | dry skin |
| | | | hypoaesthesia | body numbness |
| | | | skin burning sensation | skin burning sensation |
| | | erythemas | erythema | redness |
| | | | | redness of face |
| | | | | redness of legs |

| System Organ Class | High Level Group Term | High Level Term | Preferred Term | Low Level Term |
|---|---|---|---|---|
| | | papulosquamous conditions | oral lichen planus | oral lichen planus |
| | | pruritus NEC | pruritus | generalized itching |
| | | | | itching |
| | | pustular conditions | rash pustular | pustular rash |
| | | rashes, eruptions and exanthems NEC | rash | facial rash |
| | | | | skin rash |
| | | | rash erythematous | red rash |
| | | | rash macular | red spotty rash |
| | | | rash pruritic | itchy rash |
| | skin and subcutaneous tissue infections and infestations | skin and subcutaneous tissue bacterial infections | furuncle | boil |
| | | skin and subcutaneous tissue viral infections | herpes zoster | herpes zoster |
| | skin appendage conditions | alopecias | alopecia | hair loss |
| | | pilar disorders NEC | piloerection | goose bumps |
| vascular disorders | decreased and nonspecific blood pressure disorders and shock | circulatory collapse and shock | dizziness | dizziness |
| | | | | light headedness |
| | | vascular hypotensive disorders | hypotension | low blood pressure |

| System Organ Class | High Level Group Term | High Level Term | Preferred Term | Low Level Term |
|---|---|---|---|---|
| | vascular hypertensive disorders | vascular hypertensive disorders NEC | | hypertension |

**Supplement 3.** Factors associated with adverse effect development after SARS-CoV2 vaccination

| Characteristic | OR (95%CI) | p-value |
|---|---|---|
| **Age <65 years (vs. ≥65 years)** | 1.65 (0.96 to 2.9) | 0.072 |
| **Female sex (vs male)** | 4.05 (2.33 to 7.3) | <0.001 |
| **Vaccine** | | |
| BNT162b2 | 1 [Reference] | |
| mRNA-1273 | 2.38 (1.22 to 4.8) | 0.013 |
| JNJ-78436735 | 0.68 (0.36 to 1.3) | 0.217 |
| **Education** | | |
| None or mandatory school | 1 [Reference] | |
| Vocational training or specialized baccalaureate | 5.20 (1.56 to 17.5) | 0.007 |
| Higher education | 6.26 (1.86 to 21.2) | 0.003 |
| **Any preexisting condition (vs. none)** | 0.94 (0.51 to 1.8) | 0.848 |
| **Low opinion of vaccination (vs. high)** | 0.98 (0.46 to 2.2) | 0.960 |
| **SARS-CoV-2 Infection prior to baseline (self-reported infection or tested seropositive) (vs. none)** | 1.82 (0.79 to 4.8) | 0.188 |

**Supplement 4.** Frequency of systemic and local adverse effects reported by participants after the first and second BNT162b2 and mRNA-1273 dose.

| **Adverse Effects by Vaccine Type** | **First dose** | **Second dose** | **Overall** |
|---|---|---|---|
| **mRNA-1273** | **(N=452)** | **(N=587)** | **(N=1039)** |
| Local adverse effects | 185 (40.9%) | 129 (22.0%) | 314 (30.2%) |
| Systemic adverse effects | 267 (59.1%) | 458 (78.0%) | 725 (69.8%) |
| **BNT162b2** | **(N=319)** | **(N=378** | **(N=697)** |
| Local adverse effects | 115 (36.1%) | 86 (22.8%) | 201 (28.8%) |
| Systemic adverse effects | 204 (63.9%) | 292 (77.2%) | 496 (71.2%) |

**Supplement 5.** Self-reported duration of adverse effects

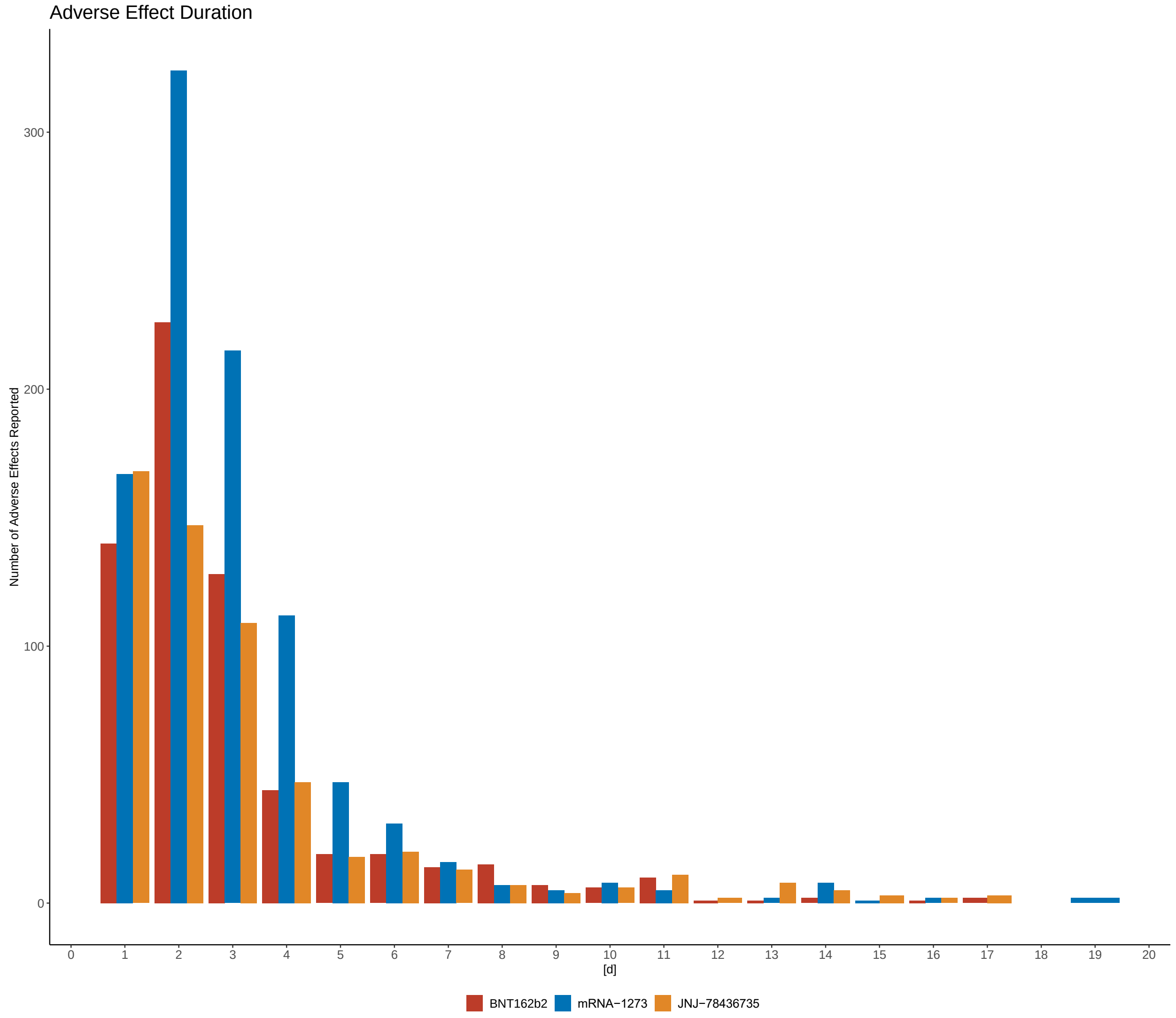


**Supplement 6.** Frequency of adverse effects according to MedDRA high level terms (NEC = not elsewhere classified)

| MedDRA High Level Terms | BNT162b2<br>(N=697) | mRNA-1273<br>(N=1039) | JNJ-78436735<br>(N=661) | Overall<br>(N=2397) |
|---|---|---|---|---|
| amnestic symptoms | 1 (0.1%) | 0 (0.0%) | 0 (0.0%) | 1 (0.0%) |
| asthenic conditions | 115 (16.5%) | 132 (12.7%) | 72 (10.9%) | 319 (13.3%) |

| | | | | |
|---|---|---|---|---|
| bone disorders NEC | 1 (0.1%) | 0 (0.0%) | 0 (0.0%) | 1 (0.0%) |
| bone related signs and symptoms | 1 (0.1%) | 0 (0.0%) | 0 (0.0%) | 1 (0.0%) |
| breathing abnormalities | 5 (0.7%) | 2 (0.2%) | 5 (0.8%) | 12 (0.5%) |
| bullous conditions | 2 (0.3%) | 0 (0.0%) | 0 (0.0%) | 2 (0.1%) |
| cardiac signs and symptoms NEC | 2 (0.3%) | 3 (0.3%) | 1 (0.2%) | 6 (0.3%) |
| circulatory collapse and shock | 2 (0.3%) | 7 (0.7%) | 9 (1.4%) | 18 (0.8%) |
| coughing and associated symptoms | 7 (1.0%) | 7 (0.7%) | 9 (1.4%) | 23 (1.0%) |
| dermal and epidermal conditions NEC | 2 (0.3%) | 0 (0.0%) | 0 (0.0%) | 2 (0.1%) |
| diarrhoea (excl infective) | 15 (2.2%) | 10 (1.0%) | 9 (1.4%) | 34 (1.4%) |
| disturbances in consciousness NEC | 1 (0.1%) | 0 (0.0%) | 0 (0.0%) | 1 (0.0%) |
| disturbances in initiating and maintaining sleep | 1 (0.1%) | 1 (0.1%) | 0 (0.0%) | 2 (0.1%) |
| dyspeptic signs and symptoms | 2 (0.3%) | 3 (0.3%) | 3 (0.5%) | 8 (0.3%) |
| dyssomnias | 3 (0.4%) | 2 (0.2%) | 5 (0.8%) | 10 (0.4%) |
| ear disorders NEC | 2 (0.3%) | 5 (0.5%) | 3 (0.5%) | 10 (0.4%) |
| erythemas | 1 (0.1%) | 3 (0.3%) | 0 (0.0%) | 4 (0.2%) |
| febrile disorders | 20 (2.9%) | 63 (6.1%) | 55 (8.3%) | 138 (5.8%) |
| feelings and sensations NEC | 23 (3.3%) | 54 (5.2%) | 53 (8.0%) | 130 (5.4%) |
| gastric and gastroenteric infections | 1 (0.1%) | 0 (0.0%) | 0 (0.0%) | 1 (0.0%) |
| gastrointestinal and abdominal pains (excl oral and throat) | 5 (0.7%) | 8 (0.8%) | 1 (0.2%) | 14 (0.6%) |
| gastrointestinal signs and symptoms NEC | 3 (0.4%) | 5 (0.5%) | 2 (0.3%) | 10 (0.4%) |
| general signs and symptoms NEC | 14 (2.0%) | 21 (2.0%) | 31 (4.7%) | 66 (2.8%) |
| gingival disorders, signs, and symptoms NEC | 1 (0.1%) | 0 (0.0%) | 1 (0.2%) | 2 (0.1%) |
| headaches NEC | 78 (11.2%) | 105 (10.1%) | 88 (13.3%) | 271 (11.3%) |

| | | | | |
|---|---|---|---|---|
| herpes viral infections | 4 (0.6%) | 4 (0.4%) | 7 (1.1%) | 15 (0.6%) |
| injection site reactions | 1 (0.1%) | 35 (3.4%) | 2 (0.3%) | 38 (1.6%) |
| joint related signs and symptoms | 13 (1.9%) | 17 (1.6%) | 15 (2.3%) | 45 (1.9%) |
| lacrimation disorders | 1 (0.1%) | 1 (0.1%) | 0 (0.0%) | 2 (0.1%) |
| lid, lash and lacrimal infections, irritations, and inflammations | 1 (0.1%) | 0 (0.0%) | 0 (0.0%) | 1 (0.0%) |
| lymphatic system disorders NEC | 5 (0.7%) | 10 (1.0%) | 8 (1.2%) | 23 (1.0%) |
| menstrual and uterine bleeding NEC | 1 (0.1%) | 2 (0.2%) | 0 (0.0%) | 3 (0.1%) |
| menstruation with increased bleeding | 7 (1.0%) | 3 (0.3%) | 3 (0.5%) | 13 (0.5%) |
| migraine headaches | 1 (0.1%) | 4 (0.4%) | 6 (0.9%) | 11 (0.5%) |
| mood alterations with depressive symptoms | 3 (0.4%) | 1 (0.1%) | 2 (0.3%) | 6 (0.3%) |
| muscle pains | 7 (1.0%) | 21 (2.0%) | 12 (1.8%) | 40 (1.7%) |
| muscle related signs and symptoms NEC | 6 (0.9%) | 2 (0.2%) | 5 (0.8%) | 13 (0.5%) |
| musculoskeletal and connective tissue conditions NEC | 5 (0.7%) | 4 (0.4%) | 1 (0.2%) | 10 (0.4%) |
| musculoskeletal and connective tissue pain and discomfort | 107 (15.4%) | 153 (14.7%) | 32 (4.8%) | 292 (12.2%) |
| nasal congestions and inflammation | 1 (0.1%) | 2 (0.2%) | 2 (0.3%) | 5 (0.2%) |
| nausea and vomiting symptoms | 10 (1.4%) | 25 (2.4%) | 16 (2.4%) | 51 (2.1%) |
| ocular disorders NEC | 2 (0.3%) | 0 (0.0%) | 0 (0.0%) | 2 (0.1%) |
| oedema NEC | 1 (0.1%) | 0 (0.0%) | 0 (0.0%) | 1 (0.0%) |
| oral dryness and saliva altered | 3 (0.4%) | 2 (0.2%) | 2 (0.3%) | 7 (0.3%) |
| osteoarthtropathies | 2 (0.3%) | 0 (0.0%) | 0 (0.0%) | 2 (0.1%) |
| pain and discomfort NEC | 34 (4.9%) | 65 (6.3%) | 42 (6.4%) | 141 (5.9%) |
| Papulosquamous conditions | 1 (0.1%) | 0 (0.0%) | 0 (0.0%) | 1 (0.0%) |

| | | | | |
|---|---|---|---|---|
| paresthesia and dysesthesia | 7 (1.0%) | 2 (0.2%) | 5 (0.8%) | 14 (0.6%) |
| pericardial disorders NEC | 1 (0.1%) | 0 (0.0%) | 0 (0.0%) | 1 (0.0%) |
| physical examination procedures and organ system status | 6 (0.9%) | 10 (1.0%) | 0 (0.0%) | 16 (0.7%) |
| pneumothorax and pleural effusions NEC | 1 (0.1%) | 0 (0.0%) | 0 (0.0%) | 1 (0.0%) |
| pruritus NEC | 2 (0.3%) | 6 (0.6%) | 2 (0.3%) | 10 (0.4%) |
| rashes, eruptions and exanthemas NEC | 3 (0.4%) | 8 (0.8%) | 7 (1.1%) | 18 (0.8%) |
| skin and subcutaneous tissue viral infections | 3 (0.4%) | 2 (0.2%) | 3 (0.5%) | 8 (0.3%) |
| supraventricular arrhythmias | 1 (0.1%) | 0 (0.0%) | 0 (0.0%) | 1 (0.0%) |
| total fluid volume increase | 1 (0.1%) | 0 (0.0%) | 0 (0.0%) | 1 (0.0%) |
| upper respiratory signs and symptoms | 2 (0.3%) | 1 (0.1%) | 2 (0.3%) | 5 (0.2%) |
| upper respiratory tract infections NEC | 7 (1.0%) | 6 (0.6%) | 7 (1.1%) | 20 (0.8%) |
| upper respiratory tract signs and symptoms | 9 (1.3%) | 12 (1.2%) | 16 (2.4%) | 37 (1.5%) |
| urticaria | 1 (0.1%) | 0 (0.0%) | 2 (0.3%) | 3 (0.1%) |
| vaccination site reactions | 113 (16.2%) | 150 (14.4%) | 52 (7.9%) | 315 (13.1%) |
| vascular hypotensive disorders | 2 (0.3%) | 2 (0.2%) | 0 (0.0%) | 4 (0.2%) |
| vertigos NEC | 13 (1.9%) | 15 (1.4%) | 16 (2.4%) | 44 (1.8%) |
| viral upper respiratory tract infections | 1 (0.1%) | 5 (0.5%) | 0 (0.0%) | 6 (0.3%) |
| allergic conditions NEC | 0 (0.0%) | 1 (0.1%) | 1 (0.2%) | 2 (0.1%) |
| alopecias | 0 (0.0%) | 2 (0.2%) | 0 (0.0%) | 2 (0.1%) |
| angioedemas | 0 (0.0%) | 2 (0.2%) | 0 (0.0%) | 2 (0.1%) |
| bladder disorders NEC | 0 (0.0%) | 1 (0.1%) | 1 (0.2%) | 2 (0.1%) |
| breast signs and symptoms | 0 (0.0%) | 1 (0.1%) | 0 (0.0%) | 1 (0.0%) |

| | | | | |
|---|---|---|---|---|
| cognitive and attention disorders and disturbances NEC | 0 (0.0%) | 3 (0.3%) | 0 (0.0%) | 3 (0.1%) |
| confusion and disorientation | 0 (0.0%) | 1 (0.1%) | 0 (0.0%) | 1 (0.0%) |
| dental pain and sensation disorders | 0 (0.0%) | 1 (0.1%) | 0 (0.0%) | 1 (0.0%) |
| depressive disorders | 0 (0.0%) | 1 (0.1%) | 2 (0.3%) | 3 (0.1%) |
| emotional and mood disturbances NEC | 0 (0.0%) | 1 (0.1%) | 0 (0.0%) | 1 (0.0%) |
| flatulence, bloating and distension | 0 (0.0%) | 1 (0.1%) | 2 (0.3%) | 3 (0.1%) |
| gingival Infections | 0 (0.0%) | 2 (0.2%) | 0 (0.0%) | 2 (0.1%) |
| heart rate and pulse investigations | 0 (0.0%) | 1 (0.1%) | 1 (0.2%) | 2 (0.1%) |
| lower respiratory tract signs and symptoms | 0 (0.0%) | 1 (0.1%) | 0 (0.0%) | 1 (0.0%) |
| muscle weakness conditions | 0 (0.0%) | 1 (0.1%) | 0 (0.0%) | 1 (0.0%) |
| myopathies | 0 (0.0%) | 1 (0.1%) | 0 (0.0%) | 1 (0.0%) |
| neurological signs and symptoms NEC | 0 (0.0%) | 4 (0.4%) | 8 (1.2%) | 12 (0.5%) |
| oral soft tissue signs and symptoms | 0 (0.0%) | 2 (0.2%) | 0 (0.0%) | 2 (0.1%) |
| pilar disorders NEC | 0 (0.0%) | 1 (0.1%) | 0 (0.0%) | 1 (0.0%) |
| pustular conditions | 0 (0.0%) | 1 (0.1%) | 0 (0.0%) | 1 (0.0%) |
| rate and rhythm disorders NEC | 0 (0.0%) | 2 (0.2%) | 2 (0.3%) | 4 (0.2%) |
| retinal structural change, deposit, and degeneration | 0 (0.0%) | 1 (0.1%) | 0 (0.0%) | 1 (0.0%) |
| sleep disorder NEC | 0 (0.0%) | 2 (0.2%) | 0 (0.0%) | 2 (0.1%) |
| tongue signs and symptoms | 0 (0.0%) | 1 (0.1%) | 0 (0.0%) | 1 (0.0%) |
| visual disorders NEC | 0 (0.0%) | 1 (0.1%) | 0 (0.0%) | 1 (0.0%) |
| visual impairment and blindness (excluding color blindness) | 0 (0.0%) | 1 (0.1%) | 2 (0.3%) | 3 (0.1%) |
| vulvovaginal disorders NEC | 0 (0.0%) | 1 (0.1%) | 0 (0.0%) | 1 (0.0%) |
| auditory nerve disorders | 0 (0.0%) | 0 (0.0%) | 2 (0.3%) | 2 (0.1%) |

| | | | | |
|---|---|---|---|---|
| bladder and urethral symptoms | 0 (0.0%) | 0 (0.0%) | 4 (0.6%) | 4 (0.2%) |
| bladder infections and inflammations | 0 (0.0%) | 0 (0.0%) | 1 (0.2%) | 1 (0.0%) |
| conjunctival infections, irritations, and inflammations | 0 (0.0%) | 0 (0.0%) | 1 (0.2%) | 1 (0.0%) |
| haemorrhoids and gastrointestinal varices (excluding oesophageal) | 0 (0.0%) | 0 (0.0%) | 1 (0.2%) | 1 (0.0%) |
| meningitis NEC | 0 (0.0%) | 0 (0.0%) | 1 (0.2%) | 1 (0.0%) |
| nasal disorders NEC | 0 (0.0%) | 0 (0.0%) | 7 (1.1%) | 7 (0.3%) |
| ocular infections, inflammations, and associated manifestations | 0 (0.0%) | 0 (0.0%) | 5 (0.8%) | 5 (0.2%) |
| olfactory nerve disorders | 0 (0.0%) | 0 (0.0%) | 1 (0.2%) | 1 (0.0%) |
| sensory abnormalities NEC | 0 (0.0%) | 0 (0.0%) | 1 (0.2%) | 1 (0.0%) |
| somatic symptom disorders | 0 (0.0%) | 0 (0.0%) | 1 (0.2%) | 1 (0.0%) |
| tendon disorders | 0 (0.0%) | 0 (0.0%) | 1 (0.2%) | 1 (0.0%) |
| tremor (excl congenital) | 0 (0.0%) | 0 (0.0%) | 1 (0.2%) | 1 (0.0%) |
| vascular hypertensive disorders NEC | 0 (0.0%) | 0 (0.0%) | 1 (0.2%) | 1 (0.0%) |
